# Higher ultraviolet light exposure is associated with lower mortality: an analysis of data from the UK Biobank cohort study

**DOI:** 10.1101/2023.07.11.23292360

**Authors:** Andrew C. Stevenson, Tom Clemens, Erola Pairo-Castineira, David J. Webb, Richard B. Weller, Chris Dibben

## Abstract

**Objective:** To examine to what extent UV exposure is associated with all-cause and cause-specific mortality.

**Design:** Prospective population-based study.

**Setting:** UK Biobank.

**Participants:** 376,729 participants with white ancestry and no missing data. Two UV exposures were assessed: sun-seeking behaviour (categorised as less active versus more active) and home latitude.

**Main outcome measures:** All-cause, cardiovascular disease (CVD), cancer and non-CVD/non-cancer mortality. Risk of residual confounding was examined using three negative control outcomes.

**Results:** The median follow-up was 12.7 years. Participants with more active sun-seeking behaviour were at a lower risk of all-cause mortality (HR=0.86; 95% confidence interval (CI) 0.80 to 0.93), CVD mortality (HR=0.81; 95% CI 0.68 to 0.95) and cancer mortality (HR=0.86; 95% CI 0.77 to 0.95) compared to participants with less active sun-seeking behaviours, adjusted for demographic, socioeconomic, behavioural and clinical confounders. More active sun seekers had around 50 extra days of survival. Participants whose home latitude was 300km farther south were also at a lower risk of all-cause mortality (HR=0.94; 95% CI 0.92 to 0.96), CVD mortality (HR=0.91; 95% CI 0.86 to 0.95) and cancer mortality (HR=0.93; 95% CI 0.90 to 0.96), adjusted for demographic, socioeconomic, behavioural and clinical confounders. Participants whose home latitude was 300km farther south had around 16 extra days of survival. Sun-seeking behaviour was not associated with two of the three negative controls and home latitude was not associated with any of the negative controls.

**Conclusions:** Greater behavioural and higher geographically related UV exposures were associated with a lower risk of all-cause, CVD and cancer mortality. This study adds to growing evidence that the benefits of UV exposure may outweigh the risks in low sunlight countries. Tailoring public health advice to both the benefits and hazards of UV exposure may reduce the burden of disease and increase life expectancy in low sunlight countries.

## INTRODUCTION

Public health messaging in the United Kingdom (UK) and other countries with a large population of European descendants has tended to emphasise the risks of ultraviolet (UV) exposure. The known association between UV radiation and melanoma pathogenesis is of particular concern.[1] Between 2006-2008 and 2016-2018, melanoma incidence increased by 32% in the UK.[2] However, mortality has remained stable over the last decade and is starting to decrease among women.[3] In 2017-2019, melanoma mortality was relatively low, representing 1% of all cancer deaths.[3] Recent evidence suggests that the benefits of UV exposure may outweigh any risks, especially in low-light environments. In a cohort of Swedish women, participants with higher levels of sun exposure lived longer than those who avoided the sun.[4] The mortality advantage was attributed to lower cardiovascular disease (CVD) and non-cancer/non-CVD mortality. In a case-control study of Swedish women with low-to-moderate sun exposure habits, women with fair phenotypes (the most UV-sensitive) had an 8% lower all-cause mortality rate than non-fair women, despite having a greater risk of dying from skin cancer.[5]

Several biologically plausible mechanisms exist for a relationship between ultraviolet A (UVA) and ultraviolet B (UVB) exposure and positive health outcomes. UVB synthesises vitamin D in exposed skin.[6] Higher levels of vitamin D have been associated with lower cancer and CVD rates in observational studies. However, recent randomised controlled trials of vitamin D supplementation and Mendelian randomisation studies do not support a causal role of vitamin D on a range of extra-skeletal health outcomes.[7-10] UVA photons have longer wavelengths and penetrate deeper into the skin.[6] Dermal UVA exposure triggers nitric oxide (NO)-mediated vasodilatation, which lowers blood pressure.[11] NO is also a negative regulator of the NLRP3 inflammasome, which is associated with a wide range of diseases, including type II diabetes and atherosclerosis.[12] New evidence suggests that UVA protects against myocardial infarction[13] and COVID-19 mortality,[14] independent of UVB.

The UK is a high latitude and low-sunlight country. The UV index, which measures the erythemal intensity of sunlight, rarely exceeds 6 (where 3-5 is classified as moderate and 6-7 high) in most parts of the UK[15, 16]. Indeed, there is a high prevalence of low vitamin D -a biomarker for low UV exposure-at the population level [17]. However, sun avoidance public health campaigns are perhaps influenced by those from extreme UV environments like Australia with pale skinned European populations.[18] Lower residential latitudes and individual sun-seeking behaviours are determinants of higher personal UV irradiation. [19, 20] This study aimed to determine to what extent UV exposure is associated with all-cause and cause-specific mortality using data from participants of the UK Biobank. We used two distinct estimates of exposures and negative control outcomes to test this question.

## METHODS

### Cohort and sample

The UK Biobank is a prospective community-based cohort of over 500,000 participants aged 37 to 73 at recruitment between 2006 and 2010, living close to 22 recruitment centres located throughout England, Wales and Scotland.[21] Each participant provided signed consent and completed a touch-screen questionnaire and computer-assisted interview on sociodemographic information, health exposures, and medical history and treatments. Participants also underwent a physical assessment and provided blood, urine and saliva samples[22]. DNA was extracted for genotyping, and 30 biomarkers were measured.[23] UK Biobank’s ethical approval was from the North West Centre for Research Ethics Committee (11/NW/0382). The present analysis was approved under the UK Biobank project 30585. Genetic skin pigmentation plays an important role in biological responses to UV exposure.[24] To limit the potential confounding effect of UV exposure, skin pigmentation and mortality, we restricted to participants of white ancestry in the present analysis using a combination of self-reported ethnic background and genetic information. Participants with missing data were excluded from the analysis.

### Patient and public involvement

No patients were involved in setting the research question or the outcome measures, nor were they involved in developing plans for design or implementation of the study. Participants of the UK Biobank can register to be updated on new results and are offered opportunities to help with further research initiatives.

### Study design

To better assess causality, we employed two design approaches; [i] two exposures were estimated originating from independent processes, allowing triangulation, and [ii] negative control outcomes were used. Triangulation involves contrasting estimates from several different aetiological approaches.[25] With different processes involved in producing the exposures, it is less likely that a single source of bias could be affecting each analysis.

We developed estimates for UV exposures derived from different social and economic contexts and we modelled these separately. By identifying different contexts, we maximise the chance that any remaining bias in our results (e.g., due to omitted variable bias) will be independent between the two models. If the two models with different biasing structures agree, there is stronger evidence that the relationship is real and not due to measurement error or bias.[26] We chose three outcomes that met the negative control criteria (that exposures of interest have no reported or plausible effect on but are subject to the same unobserved confounding as the outcomes of interest) that in particular could be affected by two confounders in our directed acyclic graph (DAG; Supplemental File 1): risky behaviours and socioeconomic factors. Any association observed between the exposure and negative control outcome indicates that there may be confounding in the main models.

The UK Biobank has data from which behavioural and geographic UV exposures can be estimated. Firstly, participants were asked ‘how many times a year would you use a solarium or sunlamp?’ As studies have shown that individuals who use solariums engage in sun-seeking behaviours[27] we recoded the responses to create a *sun-seeking behaviour* variable. We dichotomised this variable as never (‘less active sun-seeking behaviours’) or one or more times per year (‘more active sun-seeking behaviours’). Individuals who responded ‘do not know’ or ‘prefer not to answer’ were coded as missing. Secondly, the full address of each participant was used to derive a set of 1-km resolution co-ordinates, which were then used to estimate the north co-ordinate of their home location. Where full addresses were not available, co-ordinates were based on the participants’ postcode. The location co-ordinates use the Ordnance Survey (OSGB) reference system. We used this variable to calculate the *home latitude* of each participant’s residential location. For validity, we tested both these exposure measures against measured serum 25-Hydroxy Vitamin D (25(OH)D) levels. Blood samples were obtained from participants at baseline, and biological assays were performed at a central laboratory. The protocol for blood sampling has been previously described.[23]

Serum 25(OH)D, a biomarker of vitamin D, was measured by chemiluminescence immunoassay (DiaSorin Liaison XL, Italy), which has a detection range of 10 to 375 nmol/L. Several quality control procedures were performed to ensure the precision of analyses.[28] As negative controls outcomes we measured and assessed hospitalisations due to car or motorcycle accidents (ICD-10: V200-V199; V400-V499), pedestrian accidents (ICD-10: V010-V099) and cycling accidents (ICD-10: V010-V199).

### Outcomes of interest

All-cause and cause-specific mortality (CVD, cancer, and non-CVD/non-cancer death) were the primary outcomes of interest in this study. Each participant from the UK Biobank was linked to a national death registry (National Health Service (NHS) Digital or NHS Central Register) at the date of their recruitment into the study [29]. The UK Biobank receives the date of death and the primary and contributory causes, identified using the International Statistical Classification of Diseases (ICD) codes. A list of ICD-10 codes used for each cause of mortality is available in Supplementary File 2.

### Confounders

We identified several demographic, socioeconomic, and behavioural factors *a priori* (aided by our DAG) that we assume could influence both our measures of an individual’s UV exposure and mortality risk and therefore could be confounding. For the *sun-seeking behaviour* variable, we considered age at recruitment (categorised as 37-47, 48-54, 55-59, 60- 63, 64-73), sex (female or male), employment status (dichotomised as employed or unemployed), age completed full-time education (categorised as ≤15, 16 to 18, ≥19), area-level UK-adjusted index of multiple deprivation (IMD) (a continuous variable), body mass index (BMI, kg/m^2^) (categorised as <25, 25 to 30, 30+), smoking status (categorised as never, former and current), number of days a week of vigorous physical activity lasting more than 10 minutes (dichotomised as none or one or more days), north home co-ordinate (a continuous variable based on the Ordnance Survey co-ordinates), history of mental health concerns (having seen a doctor for either nerves, anxiety, tension or depression, dichotomised as yes or no), and risk-taking behaviour (dichotomised as yes or no). To ascertain risk-taking behaviour participants were asked ‘Would you describe yourself as someone who takes risks?’ We considered the same confounders for the *home latitude* variable, except for north home co-ordinate, and included sun-seeking behaviour.

Each country in the UK has a separate IMD encompassing material deprivation and other aspects such as health and crime.[30] The indices are not directly comparable because the domains, data sources and scales differ; however, they all aim to measure the same multiple deprivation concept. We assessed the range and distribution for the raw scores of the income domain, which is the same across the UK, and found them to very similar[31, 32]. Therefore, to create a UK-wide adjusted measure of IMD, we rescaled the Wales and Scotland IMDs to the distribution of the England IMD, described in the following equation:

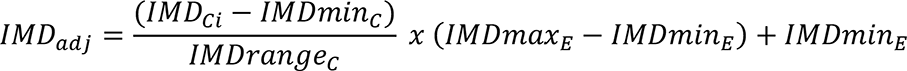

Where IMD_Ci_ is the IMD score for area i in country C; *IMDmin_c_* is the minimum IMD score in country C; *IMDrange_c_* is the difference between the minimum and maximum score in country C; IMDmax_E_ is the maximum IMD score in England and IMDmin_E_ is the minimum IMD score in England.

### Statistical analyses

Statistical analyses were performed using Stata 16 (College Station, TX: StataCorp LLC.). We calculated proportions or means and 95% confidence intervals (CIs) for each variable included in the study. Linear regression models were fitted for the UV exposures and vitamin D serum levels, adjusted for confounders identified *a priori*. The ‘predict’ command was used to estimate vitamin D serum levels at each level of the exposure assessments. Then, person-time was calculated from the date that each participant enrolled in the study to the date of death from any cause and each cause-specific death, loss to follow-up or the end of the follow-up (November 12, 2021). Age-adjusted and multivariable Cox proportional hazard regression models were fitted to estimate hazard ratios (HRs) for the binary sun-seeking behaviour and the continuous home latitude variables on all-cause and cause-specific mortality (CVD, cancer and non-cancer/non-CVD death), adjusting for confounders identified *a priori*. For the home latitude exposure, we scale the measure so that the hazard ratio represents a change of residence being 300km farther south. To estimate valid 95% CIs for time-varying HRs, nonparametric bootstrapping (300 repetitions) was used. The discriminatory ability of the models was evaluated using Harrell’s C statistic. The C-statistic ranges from 0.5 to 1.0, where 0.5 means the model performs no better than random chance and 1.0 indicates a model with perfect prediction.[33] Multivariable logistic regressions were fitted to estimate odds ratios (ORs) for sun-seeking behaviour and home latitude on the negative control outcomes (hospitalisation from car or motorcycle accidents, pedestrian accidents and cycling accidents).

We calculated the Restricted Mean Survival Time (RMST), which is a measure of the average survival time up to a certain time point.[34] We used the ’stpm2’ package to estimate the RMST, which employs a flexible parametric survival model and allows for adjustment of covariates.[35] The RMST was calculated for each exposure group separately while controlling for the same confounders identified above, and the difference in RMSTs between the groups was considered as the estimate of the difference in life expectancy at the end of the follow-up period. We estimated the crude incidence rate difference per 10,000 person-years for sun-seeking behaviour and home latitude on all cancers and cancer-specific deaths. For clarity, we considered skin cancer and the six cancer types with the highest mortality rates among the UK Biobank participants based on ICD-10 groupings. For this analysis, we recoded the home latitude variable into the northern and southern half of the UK (‘North’ vs. ‘South’) to create an exposed and unexposed group, which was necessary to calculate the incidence rate difference.

## RESULTS

There were 518,239 participants enrolled in the UK Biobank across England, Wales and Scotland, 65,106 of whom did not have white ancestry. Of those eligible, 376,729 participants had complete information for survival analyses (**Figure 1**). The total follow-up time was 4,680,913 person-years, with a median follow-up of 12.7 years. An additional 32,779 participants did not have serum 25(OH)D levels measured, leaving 343,950 participants available for the vitamin D analysis. Participant information is described in **Table 1**.

**Figure 1.**
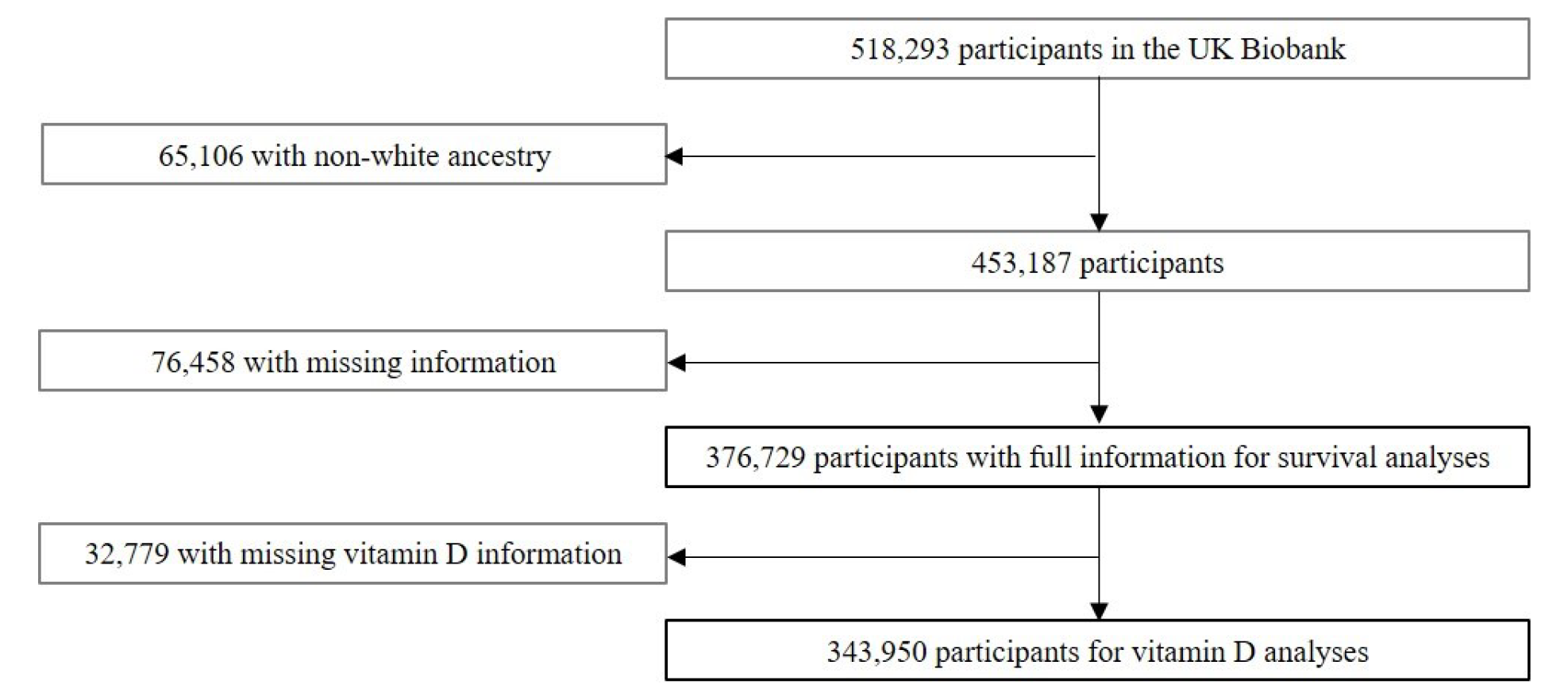
Flowchart of participant inclusion.

**Table 1.** Participant information.

| **Participant characteristics** | **All participants**<br>**(n=376,729)** | **Less active sun-**<br>**seekers**<br>**(n=358,513)** | **More active sun-**<br>**seekers**<br>**(n=18,216)** | **Highest home**<br>**latitude**<br>**(n=29,181)** | **Mid-home**<br>**latitude**<br>**(n=200,923)** | **Lowest home**<br>**latitude**<br>**(n=146,625)** |
| --- | --- | --- | --- | --- | --- | --- |
| Median follow-up time, years | 12.7 | 12.7 | 12.9 | 13.8 | 12.7 | 12.4 |
| All deaths, n | 27,111 | 26,355 | 756 | 2,703 | 15,208 | 9,200 |
| Cardiovascular disease deaths, n (%) | 5,398 (19.9) | 5,269 (20.0) | 129 (17.0) | 548 (20.3) | 3,107 (20.4) | 1,743 (18.9) |
| Cancer deaths, n (%) | 14,125 (52.1) | 13,709 (52.0) | 416 (55.0) | 1,367 (50.6) | 7,799 (51.3) | 4,959 (53.9) |
| Non-CVD/non-cancer deaths, n (%) | 7,588 (28.0) | 7,377 (28.0) | 211 (27.9) | 788 (29.2) | 4,302 (28.3) | 2,498 (27.2) |
| More active sun-seeking behaviour, % | 4.8 | 0.0 | 100.0 | 4.25 | 6.36 | 2.86 |
| Home latitude (°N), mean (95% CI) | 53.1 (53.1, 53.1) | 53.0 (53.0, 53.1) | 53.5 (53.5, 53.5) | 55.9 (55.9, 55.9) | 53.7 (53.7, 53.7) | 51.6 (51.6, 51.6) |
| Age at recruitment, % |  |  |  |  |  |  |
| 37 to 47 | 17.2 | 16.3 | 33.3 | 18.7 | 16.2 | 18.0 |
| 48 to 54 | 20.4 | 19.1 | 29.8 | 22.3 | 19.9 | 20.7 |
| 55 to 59 | 18.3 | 18.3 | 16.8 | 18.7 | 18.3 | 18.2 |
| 60 to 63 | 20.4 | 20.8 | 11.6 | 17.6 | 21.0 | 20.0 |
| 64 to 73 | 23.8 | 24.6 | 8.4 | 22.7 | 24.6 | 23.1 |
| Female, % | 53.7 | 52.9 | 68.7 | 55.0 | 52.9 | 54.5 |

| **Participant characteristics** | **All participants**<br>**(n=376,729)** | **Less active sun-**<br>**seekers**<br>**(n=358,513)** | **More active sun-**<br>**seekers**<br>**(n=18,216)** | **Highest home**<br>**latitude (**<br>**(n=29,181)** | **Mid-home**<br>**latitude**<br>**(n=200,923)** | **Lowest home**<br>**latitude**<br>**(n=146,625)** |
| --- | --- | --- | --- | --- | --- | --- |
| BMI (kg/m<sup>2</sup>), % |  |  |  |  |  |  |
| <25 | 33.6 | 33.5 | 35.9 | 34.0 | 31.3 | 36.7 |
| 25 to 30 | 42.8 | 42.9 | 42.2 | 42.8 | 43.8 | 41.5 |
| 30+ | 23.5 | 23.6 | 21.9 | 23.2 | 24.8 | 21.8 |
| Adjusted IMD,<br>mean (95% CI) | 16.2 (16.1, 16.2) | 16.0 (16.0, 16.1) | 19.6 (19.4, 19.8) | 12.8 (12.6, 13.0) | 18.1 (18.0, 18.1) | 14.2 (14.2, 14.3) |
| Smoking status,<br>% |  |  |  |  |  |  |
| Never | 54.6 | 54.9 | 49.0 | 56.2 | 54.8 | 54.0 |
| Former | 35.7 | 35.7 | 35.4 | 32.2 | 35.6 | 36.4 |
| Current | 9.8 | 9.5 | 15.7 | 11.7 | 9.6 | 9.6 |
| One or more<br>days/week of<br>vigorous physical<br>activity (10+<br>minutes), % | 62.3 | 62.0 | 69.3 | 59.9 | 61.4 | 64.1 |
| Employed, % | 58.2 | 57.2 | 78.0 | 60.2 | 55.8 | 61.1 |
| Age completed<br>education, % |  |  |  |  |  |  |
| <=15 | 20.4 | 20.5 | 19.1 | 20.7 | 25.0 | 14.1 |
| 16 to 18 | 37.8 | 37.2 | 69.4 | 31.6 | 38.6 | 38.1 |
| >=19 | 41.7 | 42.3 | 30.6 | 47.7 | 36.5 | 47.8 |
| History of mental<br>health concerns<br>(%) | 33.8 | 33.3 | 43.7 | 33.8 | 34.9 | 32.4 |
| Risk taking<br>behaviour (%) | 26.1 | 25.6 | 35.0 | 25.3 | 25.2 | 27.5 |

CVD= cardiovascular disease; BMI= body mass index; IMD = index of multiple deprivation

### UV exposures and vitamin D

Unadjusted and adjusted vitamin D levels by exposure assessment are show in **Table 2**. Vitamin D levels were higher among participants with more active sun-seeking behaviour compared to participants with less active sun-seeking behaviour after adjusting for age, sex, employment status, age completed education, adjusted IMD, BMI, smoking status, physical activity, north home co-ordinate, history of mental health concerns and risk-taking behaviour. Vitamin D levels were also higher among participants residing at lower latitudes after adjusting for age, sex, employment status, age completed education, adjusted IMD, BMI, smoking status, physical activity, history of mental health concerns, sun-seeking behaviour and risk-taking behaviour. This suggests that our two exposure measures are capturing genuine differences in UV exposure.

**Table 2.** Serum 25(OH)D levels by UV exposure. Adjusted sun-seeking behaviour models included age, sex, employment status, age completed education, adjusted IMD, BMI, smoking status, physical activity, north home co-ordinate, history of mental health concerns and risk-taking behaviour. Adjusted home latitude models also included sun-seeking behaviour.

| UV exposure | Vitamin D serum, nmol/L (95% CI) |  |
| --- | --- | --- |
|  | Unadjusted | Adjusted |
| Sun-seeking behaviour (n=343,950) |  |  |
| Less active | 49.0 (49.0, 49.1) | 48.9 (48.9, 49.0) |
| More active | 65.3 (65.0, 65.6) | 67.2 (66.9, 67.5) |
| Home latitude* (°N; n=343,950) |  |  |
| 58.1 | 43.4 (43.2, 43.7) | 43.1 (42.9, 43.4) |
| 55.4 | 46.8 (46.7, 47.0) | 46.7 (46.6, 46.8) |
| 52.7 | 50.2 (50.2, 50.3) | 50.3 (50.2, 50.3) |
| 50.0 | 53.6 (53.5, 53.8) | 53.8 (53.7, 54.0) |

UV= ultraviolet
\*each change in latitude represents a 300km change.

### Survival analyses

**Figure 2** shows the associations between sun-seeking behaviour and all-cause, CVD, cancer and non-CVD/non-cancer mortality. Multivariable models were adjusted for age, sex, employment status, age completed education, adjusted IMD, BMI, smoking status, physical activity, north home co-ordinate, history of mental health concerns and risk-taking behaviour.

**Figure 2.**
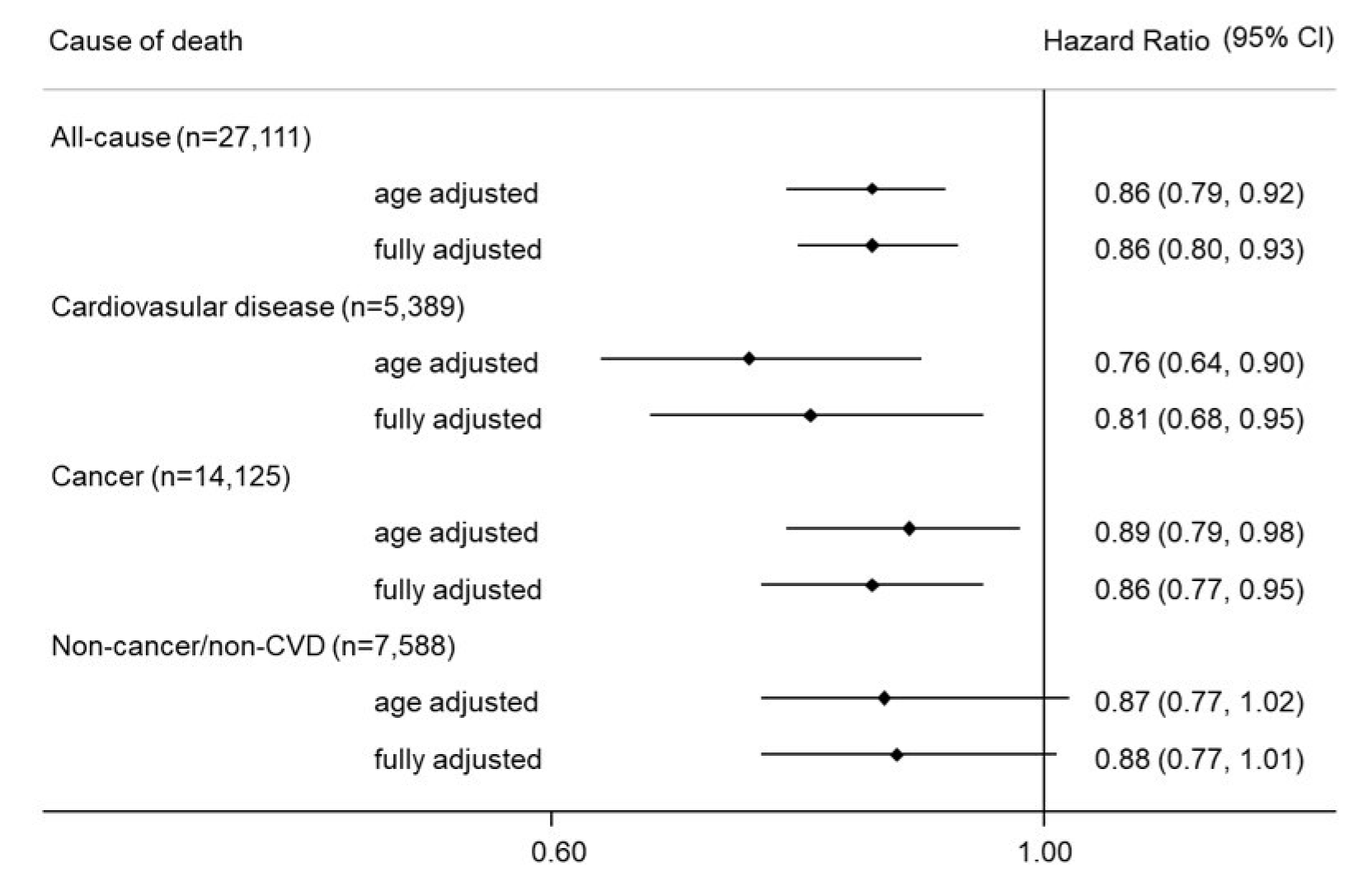
Associations between sun-seeking behaviour and mortality. Fully adjusted models included age, sex, employment status, age completed education, adjusted IMD, BMI, smoking status, physical activity, north home co-ordinate, history of mental health concerns and risk-taking behaviour CVD = cardiovascular disease

Results from the multivariable Cox regression models suggest a 14% lower risk of all-cause mortality, a 19% lower risk of CVD mortality, and a 14% lower risk of cancer mortality amongst participants with more active sun-seeking behavioural habits versus those with less active sun-seeking behaviour. Harrell’s C-statistic ranged from 0.71 for the cancer mortality model to 0.78 for the CVD mortality model. This represents the probability that, given two randomly selected individuals, the one with the higher predicted risk of death will experience the death first.[36] At the end of the follow-up period (15.6 years), life expectancy was 50 days longer for more active-sun seekers than less active sun-seekers.

**Figure 3** shows the associations between home latitude and all-cause, CVD, cancer and non-CVD/non-cancer mortality. Multivariable models were adjusted for age, sex, employment status, age completed education, adjusted IMD, BMI, smoking status, physical activity, history of mental health concerns, sun-seeking behaviour and risk-taking behaviour. Results from the multivariable Cox regression models suggest a 6% lower risk of all-cause mortality, a 9% lower risk of CVD mortality, and a 7% lower risk of cancer mortality for participants whose home latitude was 300km farther south. Harrell’s C-statistic ranged from 0.71 for the cancer mortality model to 0.79 for the CVD mortality model. At the end of the follow-up period, life expectancy was 16 days longer for participants whose home latitude was 300km farther south.

**Figure 3.**
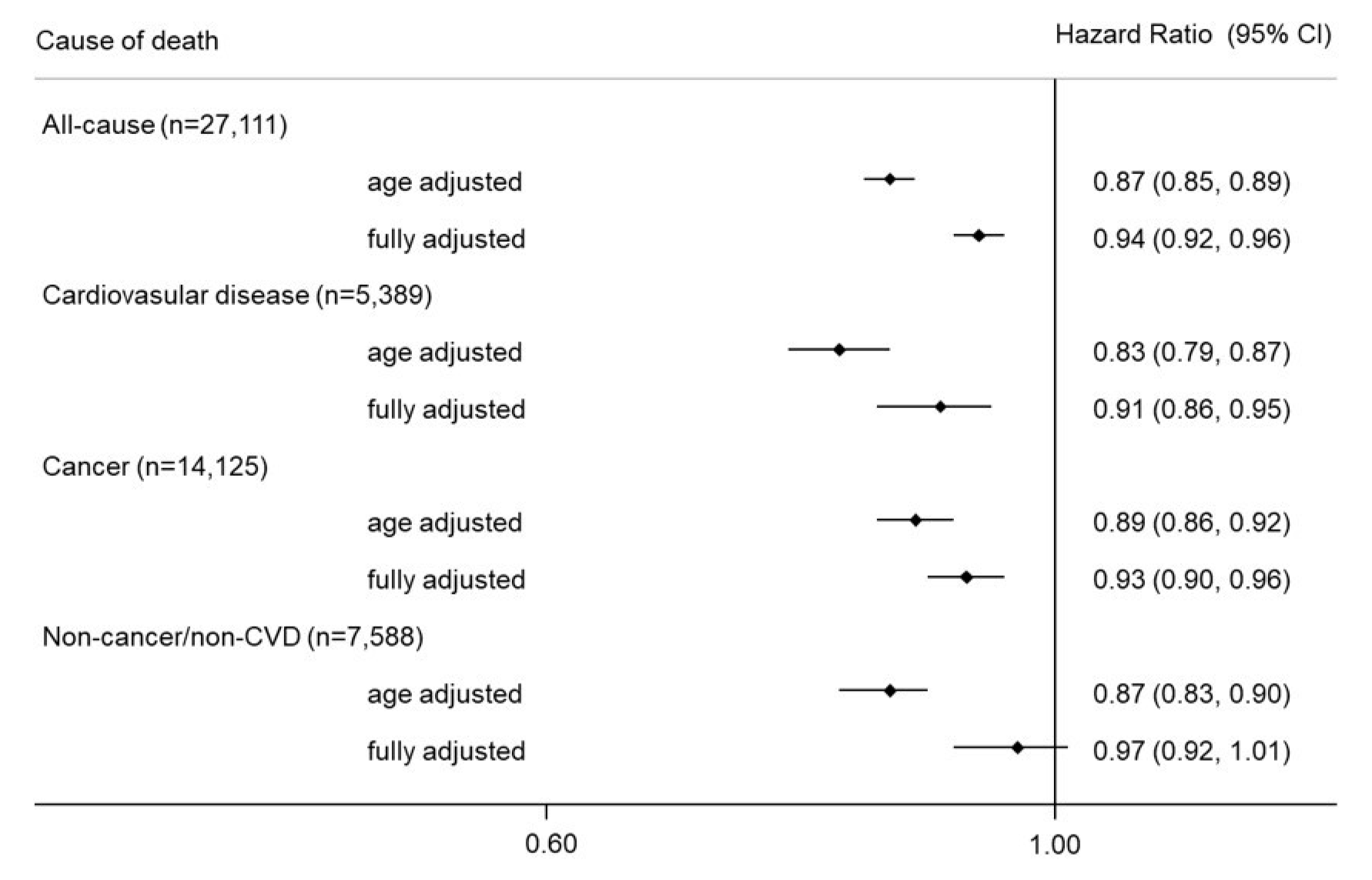
Associations between home latitude and mortality. Fully adjusted models included age, sex, employment status, age completed education, adjusted IMD, BMI, smoking status, physical activity, history of mental health concerns, sun-seeking behaviour and risk-taking behaviour. CVD = cardiovascular disease

### Negative control outcomes

In fully adjusted models, sun-seeking behaviour was not associated with car and motorcycle (OR=1.13; 95% CI 0.95, 1.34) or pedestrian hospitalisations (OR=1.00, 95% CI 0.71 to 1.41). However, more active sun-seekers had lower odds of cycling hospitalisations (OR=0.79; 95% CI 0.63 to 0.95). In fully adjusted models, home latitude was not associated with car and motorcycle (OR=1.02; 95% CI 0.95 to 1.12), pedestrian (OR=0.99; 95% CI 0.86 to 1.15) or cycling hospitalisations (OR=1.01; 95% CI 0.94 to 1.09).

### Cancer incidence rate differences

Among participants with more active sun-seeking behaviour compared to participants with less active sun-seeking behaviour, the unadjusted absolute incidence rate difference was lower for all cancer deaths, skin cancer deaths, deaths from cancers of the digestive organs, deaths from cancers of the respiratory and intrathoracic organs, cancers of lymphoid, haematopoietic and related tissue, breast cancer deaths and deaths from cancer of male genital organs. Among participants who live in the southern half of the UK versus the northern half, the unadjusted absolute incidence rate difference was lower for all cancer deaths, deaths from cancers of the digestive organs, and deaths from cancers of the respiratory and intrathoracic organs. There was no significant incidence rate difference for the remaining cancer types considered (**Figure 4**).

**Figure 4.**
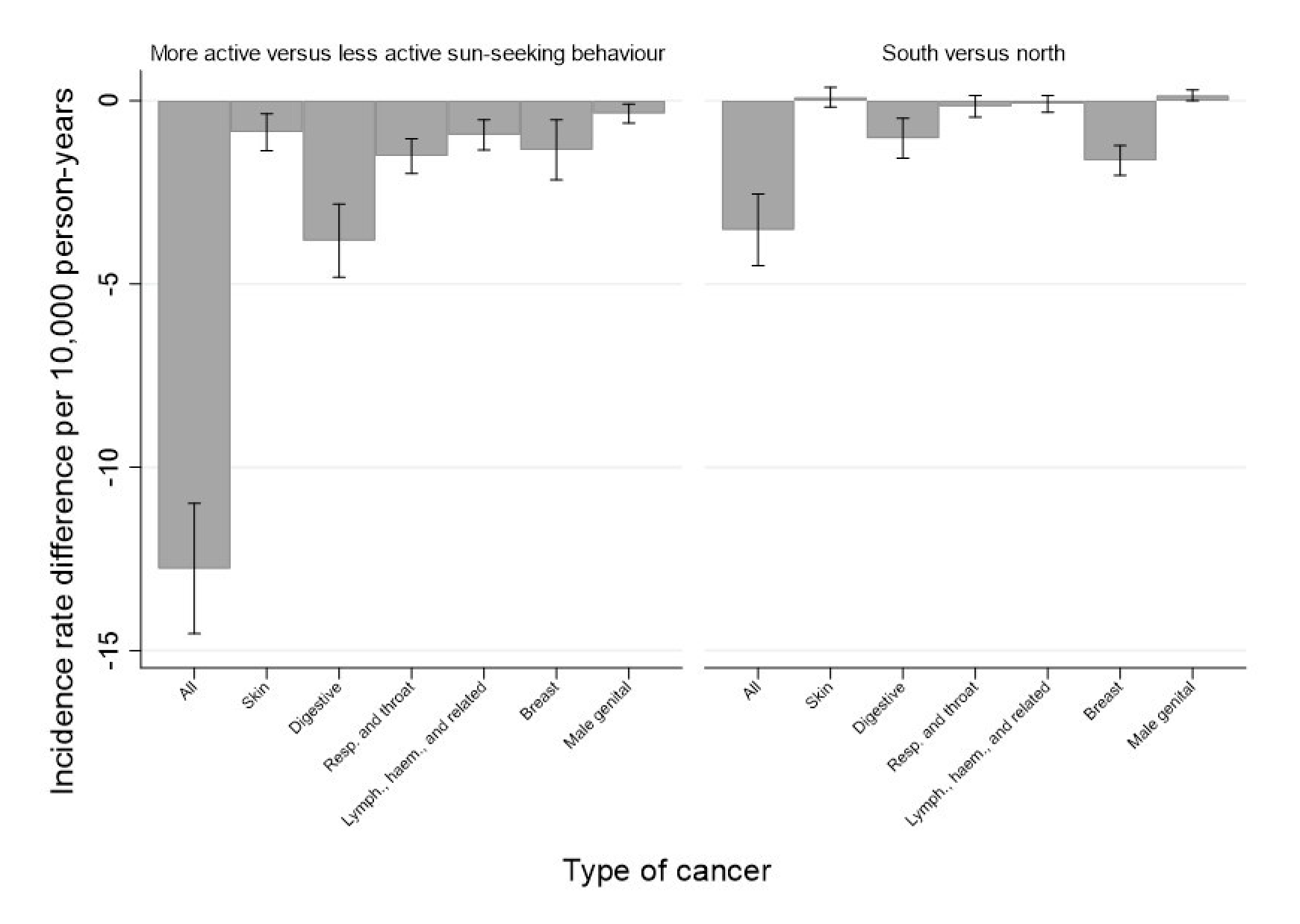
Cancer incidence rate differences for sun-seeking behaviour and home latitude. Resp.= respiratory; Lymph.= lymphoid; Haem= haematopoietic

## DISCUSSION

We find that UK Biobank participants with more active sun-seeking behaviours and who lived at lower latitudes (with a higher average UV exposure) have a lower risk of all-cause, CVD and cancer mortality. These results are consistent for two very different types of exposure, suggesting, alongside adjustment and confirmation of appropriate adjustment through negative outcome controls, that it is UV exposure and not an unmeasured variable that leads to the lower mortality risk.

These results add to the growing literature suggesting that UV exposure is associated with lower mortality risk. Results from prospective cohort studies in Sweden, at a similar latitude to the UK, demonstrated an inverse relationship between more active sun-seeking behaviours and all-cause mortality[37, 38] and CVD and non-cancer/non-CVD mortality.[4] A matched case-control analysis in a Danish study suggested that individuals with a history of non-melanoma skin cancer had a lower risk of all-cause mortality compared to the cancer-free population. [39] Several studies have also suggested an association between latitude and mortality, whereby living closer to the equator was associated with higher life expectancy, lower CVD mortality and lower mortality from several cancers.[40-44]

Participants with more active sun-seeking behaviour and those living at lower latitudes had lower crude mortality from cancers of the digestive system and breast cancer. Participants with more active sun-seeking behaviour also had lower crude mortality from skin cancer, cancers of respiratory and intrathoracic organs, cancers of lymphoid, haematopoietic and related tissue, and cancers of the male genital organs. Previous research found an inverse relationship between solar UV exposure and cancer mortality in multiple sites, including the bladder, colon, Hodgkin lymphoma, prostate, stomach, and breast.[45] In a large randomised controlled trial (the VITAL study), vitamin D supplementation was not associated with cancer incidence but there was a signal of reduced cancer mortality (HR=0.83; 95% CI 0.67 to 1.02) and a significant reduction in cancer mortality that accounted for latency by excluding the first year (HR=0.79; 95% CI 0.63 to 0.99).[46]

The links between sun exposure and melanoma development and melanoma mortality are complex. Over-diagnosis of melanoma within some health systems may well be a significant issue [47] with incidence closely linked to markers of diagnostic scrutiny but unrelated to environmental UV.[48] Our data describe the far more robust endpoint of deaths from skin cancer and is thus less subject to such errors. In the most recent WHO classification of melanoma, where aetiology and development pathway are considered, the most common form of melanoma is the Low Cumulative Sun Exposure (Low CSD) melanoma, which is analogous to Superficial Spreading Melanoma in the traditional McGovern/Clark classification.[49] As the name suggests, these melanomas are typified by the absence of signs of chronic sun exposure and predominantly occur on intermittently sun exposed body sites. Most melanoma is thus a disease of intermittent burning sun exposure, particularly in childhood. Outdoor workers have no increase in melanoma incidence compared to indoor workers.[50] Multiple studies have correlated higher vitamin D levels -a biomarker for chronic sun exposure-with reduced melanoma mortality.[51] Evidence suggests that patients with melanoma have an increased but low risk of melanoma mortality and live longer than people in the general population.[52] Studying the relationship between UV exposure and observed melanoma incidence may not be a good indicator of the relationship between UV exposure and melanoma mortality.

It is commonly hypothesised that UVB-mediated vitamin D production is the causal mechanism between exposure to sunlight and better health outcomes. Evidence from observational studies indicates inverse associations of vitamin D with risks of mortality from cardiovascular disease, cancer and other causes.[53] However, several Mendelian randomisation studies and clinical trials do not support the beneficial role of vitamin D and vitamin D supplementation on several extra-skeletal health outcomes.[7-10] A recent review of several clinical trials found that providing vitamin D supplementation to adults who are vitamin D-replete did not prevent cancer, CVD events, or the progression of type 2 diabetes.[54]

UVB rays may provide different health benefits from vitamin D supplements as UVB radiation has been shown to activate the central neuroendocrine system to regulate global homeostasis independent of vitamin D synthesis[55]. A causal role of UVA in reducing the risk of mortality is also biologically plausible. UVA exposure mobilises NO reserves in the skin and causes vasodilatation,[56] which reduces blood pressure.[11] High blood pressure is a risk factor for death and CVD.[57] NO also regulates the NLRP3 inflammasome, which plays a key role in the inflammatory response.[12] Dysfunction of the NLRP3 inflammasome can contribute to chronic inflammation, which is a key feature for the development and progression of many cancers and is associated with cardiovascular disease, metabolic disorders and infections.[58]

A strength of our study is that it used a large sample of individuals followed up over time. Participants were linked to mortality registry data, which minimises the potential for measurement error and enhances the accuracy of mortality outcomes. We used multiple UV exposures with different confounding structures and found similar patterns of protection from mortality, suggesting that the relationships are not spurious. We also included three different negative control outcomes with independent confounding structures (hospitalisation from car and motorcycle accidents, pedestrian accidents and cycling accidents). There was no association between the negative control outcomes and home latitude, suggesting that unmeasured confounding is not biasing the results. Similarly, sun-seeking behaviour was not associated with hospitalisation from car and motorcycle accidents and pedestrian accidents. However, more active sun-seekers had lower odds of being hospitalised from a cycling accident. This might indicate unmeasured confounding but might also be due to this negative outcome not working well. The finding could be a result of more active sun-seekers being less likely to cycle (a mediator of the association), which may be driving this protective association.

There are several limitations to our study. The results are based on observational data, which may suffer from residual confounding. Additionally, UK Biobank participants are not representative of the UK population and there is evidence of a healthy volunteer selection bias.[59] However, representativeness is not necessary for causal inference.[60] Selection can induce collider bias in cohort studies, whereby participation is influenced by the exposure and the outcome, leading to biased estimates of associations.[61] However, sun-seeking behaviour and home latitude do not likely influence participation or retention into the cohort to a large degree, especially compared to other exposures such as reduced cognitive ability or having severe illness. The follow-up time was relatively short, reducing the number of deaths and therefore the power of the study. Our sun-seeking behaviour variable was collected through a questionnaire, which asked respondents how many times a year they use a solarium or sun lamp. Reporting bias and social desirability bias are a concern, as participants may not have accurately recalled their use of solariums and sun lamps, or they may not want to report behaviour often deemed as unhealthy. This variable may not capture other sun-seeking behaviours but solarium users are known to exhibit sun-seeking behaviour[27]. Measured vitamin D levels support this behavioural finding in our cohort. The home latitude variable does not capture travel beyond their residential location, which could lead to variation in UV exposure. Despite this, there was a dose-response relationship between lower home latitudes and vitamin D, indicating that lower home latitude is a determinant of higher UV exposure. Another limitation is that information was collected from participants during their baseline assessment visits. Participants’ behaviour may have changed throughout the study, potentially introducing misclassification of the exposures and confounding variables over the follow-up period.

Current public health messaging emphasises the hazards of UV exposure in relation to skin cancer development. However, our study adds to growing evidence that the benefits of UV exposure on mortality outweigh the risks in low sunlight environments. Tailoring public health advice to weigh both hazards and benefits of UV exposure may reduce the burden of disease and increase life expectancy in the population countries with low sunlight. Policy agendas focusing on designing neighbourhoods to promote active living (e.g., the 20-minute neighbourhood concept whereby residents can reach amenities within short, non-motorised trip, from home[62]) may synergistically benefit population health through increased physical activity and higher UV exposure. Future studies that investigate the independent effects of UVA and UVB exposure on health outcomes and clinical trials of personal UV lamp use are warranted.

## Data Availability

Data is not available

