## Supplemental File 1 for "Higher ultraviolet light exposure is associated with lower mortality: an analysis of data from the UK Biobank cohort study"

**Supplementary file 1.** Directed Acyclic Graphs illustrating hypothesized relationships between variables.

A. Sun-seeking exposure and mortality outcomes

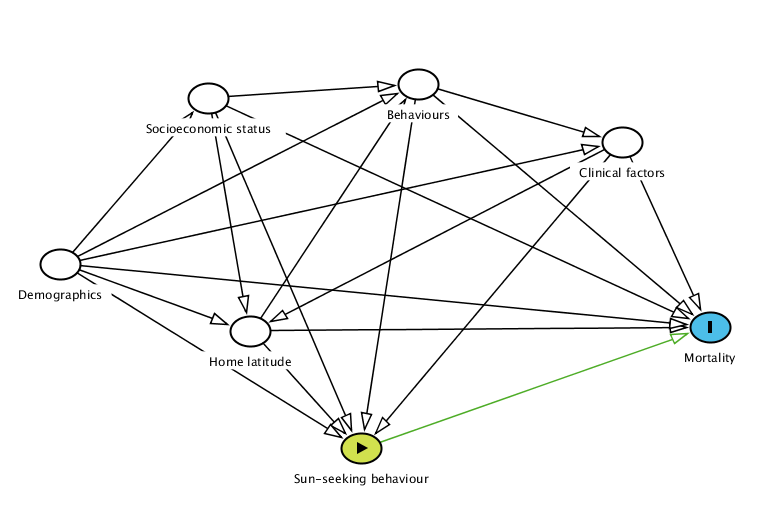

Demographics: age and sex; Socioeconomic status: employment, area-level deprivation and education; Behaviours: smoking, physical activity and risk taking; Clinical factors: body mass index

B. Home latitude exposure and mortality outcomes

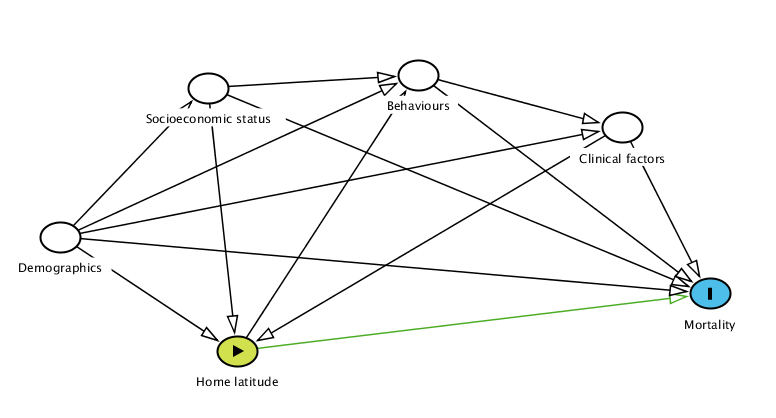

Demographics: age and sex; Socioeconomic status: employment, area-level deprivation and education; Behaviours: smoking, physical activity, risk taking and sun-seeking behaviour; Clinical factors: body mass index
