## Supplemental file 2 for "Higher ultraviolet light exposure is associated with lower mortality: an analysis of data from the UK Biobank cohort study"

**Supplementary File 2.** International Classification of Diseases (ICD)-10 codes for cardiovascular disease (CVD), cancer and non-CVD/non-cancer deaths.

**Cardiovascular disease death**

I05-I09 Chronic rheumatic heart diseases

I10-I15 Hypertensive diseases

I20-I25 Ischaemic heart diseases

I26-I28 Pulmonary heart disease and diseases of pulmonary circulation

I30-I52 Other forms of heart disease

I60-I69 Cerebrovascular diseases

I70-I79 Diseases of arteries, arterioles and capillaries

I80-I89 Diseases of veins, lymphatic vessels and lymph nodes, not elsewhere classified

**Cancer death ICD-10 codes**

C00-C14 Malignant neoplasms of lip, oral cavity and pharynx

C15-C26 Malignant neoplasms of digestive organs

C30-C39 Malignant neoplasms of respiratory and intrathoracic organs

C40-C41 Malignant neoplasms of bone and articular cartilage

C43-C44 Melanoma and other malignant neoplasms of skin Skin cancer

C45-C49 Malignant neoplasms of mesothelial and soft tissue

C50-C50 Malignant neoplasm of breast

C51-C58 Malignant neoplasms of female genital organs

C60-C63 Malignant neoplasms of male genital organs

C64-C68 Malignant neoplasms of urinary tract

C69-C72 Malignant neoplasms of eye, brain and other parts of central nervous system

C73-C75 Malignant neoplasms of thyroid and other endocrine glands

C76-C80 Malignant neoplasms of ill-defined, secondary and unspecified sites

C81-C96 Malignant neoplasms, stated or presumed to be primary, of lymphoid, haematopoietic and related tissue

C97-C97 Malignant neoplasms of independent (primary) multiple sites

D10-D36 Benign neoplasms

**Non-CVD/non-cancer death**

A00-A09 Intestinal infectious diseases62

A15-A19 Tuberculosis6

A20-A28 Certain zoonotic bacterial diseases2

A30-A49 Other bacterial diseases176

A80-A89 Viral infections of the central nervous system55

A92-A99 Arthropod-borne viral fevers and viral haemorrhagic fevers1

B00-B09 Viral infections characterized by skin and mucous membrane lesions

B15-B19 Viral hepatitis

B20-B24 Human immunodeficiency virus [HIV] disease

B25-B34 Other viral diseases

B35-B49 Mycoses

B50-B64 Protozoal diseases

B90-B94 Sequelae of infectious and parasitic diseases

B99-B99 Other infectious diseases

D55-D59 Haemolytic anaemias

D60-D64 Aplastic and other anaemias

D65-D69 Coagulation defects, purpura and other haemorrhagic conditions

D70-D77 Other diseases of blood and blood-forming organs

D80-D89 Certain disorders involving the immune mechanism

E00-E07 Disorders of thyroid gland

E10-E14 Diabetes mellitus

E15-E16 Other disorders of glucose regulation and pancreatic internal secretion

E20-E35 Disorders of other endocrine glands

E40-E46 Malnutrition

E65-E68 Obesity and other hyperalimentation

E70-E90 Metabolic disorders

F00-F09 Organic, including symptomatic, mental disorders

F10-F19 Mental and behavioural disorders due to psychoactive substance use

F20-F29 Schizophrenia, schizotypal and delusional disorders

F30-F39 Mood [affective] disorders

F50-F59 Behavioural syndromes associated with physiological disturbances and physical factors

F80-F89 Disorders of psychological development

G00-G09 Inflammatory diseases of the central nervous system

G10-G14 Systemic atrophies primarily affecting the central nervous system

G20-G26 Extrapyramidal and movement disorders

G30-G32 Other degenerative diseases of the nervous system

G35-G37 Demyelinating diseases of the central nervous system

G40-G47 Episodic and paroxysmal disorders

G50-G59 Nerve, nerve root and plexus disorders

G60-G64 Polyneuropathies and other disorders of the peripheral nervous system

G70-G73 Diseases of myoneural junction and muscle

G80-G83 Cerebral palsy and other paralytic syndromes

G90-G99 Other disorders of the nervous system

H65-H75 Diseases of middle ear and mastoid

J00-J06 Acute upper respiratory infections

J09-J18 Influenza and pneumonia

J20-J22 Other acute lower respiratory infections

J30-J39 Other diseases of upper respiratory tract

J40-J47 Chronic lower respiratory diseases

J60-J70 Lung diseases due to external agents

J80-J84 Other respiratory diseases principally affecting the interstitium

J85-J86 Suppurative and necrotic conditions of lower respiratory tract

J90-J94 Other diseases of pleura

J95-J99 Other diseases of the respiratory system

K00-K14 Diseases of oral cavity, salivary glands and jaws

K20-K31 Diseases of oesophagus, stomach and duodenum

K35-K38 Diseases of appendix

K40-K46 Hernia

K50-K52 Noninfective enteritis and colitis

K55-K64 Other diseases of intestines

K65-K67 Diseases of peritoneum

K70-K77 Diseases of liver

K80-K87 Disorders of gallbladder, biliary tract and pancreas

K90-K93 Other diseases of the digestive system

L00-L08 Infections of the skin and subcutaneous tissue

L10-L14 Bullous disorders

L40-L45 Papulosquamous disorders

L50-L54 Urticaria and erythema

L80-L99 Other disorders of the skin and subcutaneous tissue

M00-M03 Infectious arthropathies

M05-M14 Inflammatory polyarthropathies

M15-M19 Arthrosis

M20-M25 Other joint disorders

M30-M36 Systemic connective tissue disorders

M40-M43 Deforming dorsopathies

M45-M49 Spondylopathies

M60-M63 Disorders of muscles

M70-M79 Other soft tissue disorders

M80-M85 Disorders of bone density and structure

M86-M90 Other osteopathies

N00-N08 Glomerular diseases

N10-N16 Renal tubulo-interstitial diseases

N17-N19 Renal failure

N20-N23 Urolithiasis

N25-N29 Other disorders of kidney and ureter

N30-N39 Other diseases of urinary system

N40-N51 Diseases of male genital organs

N60-N64 Disorders of breast

N70-N77 Inflammatory diseases of female pelvic organs

N80-N98 Noninflammatory disorders of female genital tract

O30-O48 Maternal care related to the fetus and amniotic cavity and possible delivery problems

Q00-Q07 Congenital malformations of the nervous system

Q20-Q28 Congenital malformations of the circulatory system

Q38-Q45 Other congenital malformations of the digestive system

Q60-Q64 Congenital malformations of the urinary system

Q65-Q79 Congenital malformations and deformations of the musculoskeletal system

Q80-Q89 Other congenital malformations

Q90-Q99 Chromosomal abnormalities, not elsewhere classified

R00-R09 Symptoms and signs involving the circulatory and respiratory systems

R47-R49 Symptoms and signs involving speech and voice

R50-R69 General symptoms and signs

R95-R99 Ill-defined and unknown causes of mortality

V01-V09 Pedestrian injured in transport accident

V10-V19 Pedal cyclist injured in transport accident

V20-V29 Motorcycle rider injured in transport accident

V40-V49 Car occupant injured in transport accident

V80-V89 Other land transport accidents

V90-V94 Water transport accidents

V95-V97 Air and space transport accidents

W00-W19 Falls

W20-W49 Exposure to inanimate mechanical forces

W50-W64 Exposure to animate mechanical forces

W65-W74 Accidental drowning and submersion

W75-W84 Other accidental threats to breathing

W85-W99 Exposure to electric current, radiation and extreme ambient air temperature and pressure

X00-X09 Exposure to smoke, fire and flames

X10-X19 Contact with heat and hot substances

X20-X29 Contact with venomous animals and plants

X30-X39 Exposure to forces of nature

X40-X49 Accidental poisoning by and exposure to noxious substances

X58-X59 Accidental exposure to other and unspecified factors

X60-X84 Intentional self-harm

X85-X99 Assault

Y10-Y34 Event of undetermined intent

Y40-Y59 Drugs, medicaments and biological substances causing adverse effects in therapeutic use

Y60-Y69 Misadventures to patients during surgical and medical care

Y83-Y84 Surgical and other medical procedures as the cause of abnormal reaction of the patient, or of later complication, without mention of misadventure at the time of the procedure

Y85-Y89 Sequelae of external causes of morbidity and mortality

U00-U49 Provisional assignment of new diseases of uncertain etiology or emergency use
